# Assessing Antimicrobial Use Patterns in Christian Health Association of Malawi (CHAM) Health Facilities: A Cross-Sectional Study Protocol

**DOI:** 10.1101/2024.06.22.24309277

**Authors:** Evans Chimayi Chirambo, Dumisani Nkhoma, Francis Chiumia, Sharon Odeo, Patience Khomani, Innocent Chibwe, Simon Matchado, Pacharo Matchere, Beatrice Chiweza, Elled Mwenyekonde, Elizabeth Kampira, Collins Mitambo, Happy Makala

## Abstract

**Introduction:** The threat of antimicrobial resistance (AMR) in Malawi is high with reported mortality of 19,300 annually, the 23rd highest age-standardised mortality. One of the drivers of AMR is misuse of antibiotics, a phenomenon that has not been adequately researched in Malawi. This study aims to investigate antimicrobial use patterns using prescribing, patient and facility indicators Christian Health Association of Malawi (CHAM) health facilities.

**Methods:** This will be a multiple cross-sectional study which will collect data from facility (194), patients (388), and prescriptions (3,104). Data will be collected using KoboToolbox v2021, and exported into Microsoft Excel version 2016 for cleaning and coding. Variables will be categorized according to the antimicrobial use indicators. The study will use STATA Version 14 statistical software for data analysis. Subsequently, facilities will be put in GPS map to show hotspots of irrational antimicrobial use. The study will run from January 2024 to December 2025.

**Discussion:** This study will provide detailed information on frequently used antimicrobials, the cost of antimicrobials relative to medicine budget, the intensity of exposure to antimicrobials, the availability of antimicrobials, patients’ understanding of antimicrobials use, and availability of important documents for antimicrobial use. Secondarily, the study will unravel the prevalence of irrational antimicrobial use, the main factors contributing to it, and location where irrational use is most prevalent. These findings will inform the national antimicrobial stewardship action plan, aiming to safeguard the available antimicrobials.

**Ethics and Dissemination:** The protocol has been approved by Malawi’s College of Medicine Research Ethics Committee (COMREC) [Protocol # P.04/24-0651]. Participation in the study will be voluntary, and consent and assent will be sought during data collection. Data will be handled confidentially, with findings disseminated in conferences, to key AMR focused stakeholders, and finally through manuscript publication in a peer reviewed journal.

## Introduction

Antimicrobial resistance (AMR) has emerged as a global health crisis, posing a significant threat to the effective treatment of infectious diseases. World Health Organisation (WHO) estimates that 1.27 million annual deaths are directly linked to bacterial AMR while 4.97 million annual deaths are associated with bacterial AMR (1). It is further predicted that the global annual mortality rate will reach 10 million by 2050 if no measures are taken to curb AMR (2). In Malawi, 3,600 deaths are directly caused by bacterial AMR annually and 15,700 deaths associated with bacterial AMR, the 23rd highest age-standadised mortality rate per 100,000 population associated with AMR across 204 countries (3). Unfortunately, drivers of AMR such as irrational antimicrobial use are not well elucidated in Malawi.

Christian Health Association of Malawi (CHAM), established in 1966, is a faith-based non-governmental organization which runs 194 health facilities. These health facilities are categorized as full-fledged hospitals which are similar to a district hospital and offer specialised healthcare services, community hospital which are mid-level hospitals with very few or no specialised healthcare services, and health centres which provide basic healthcare services.The health facilities are distributed in 27 of 28 Malawi’s political districts except Mwanza District, and offers up to 30% of the national healthcare services, second only to the State, with more focus on poor people in hard-to-reach areas (4).

CHAM is one of the highest consumers of antimicrobials in Malawi. However, data is lacking on antimicrobial use in CHAM’s health facilities. Specifically, no study has looked at antimicrobial use patterns such as prescribing indicators, facility indicators and patient indicators for any of CHAM’s health facilities. This study aims to investigate antimicrobial use in CHAM health facilities using facility, patient and prescribing indicators. Understanding the patterns of antimicrobial use within CHAM will facilitate the development of targeted interventions to promote appropriate prescribing and facility practices and optimize patient outcomes.

## Objectives

### The broad objective

To investigate whether antimicrobials are properly used using prescribing, facility and patient indicators.

### The specific objectives

1. To find out the extent of antimicrobial use based on prescribing, facility and patient indicators.
2. To measure the prevalence of irrational use of antimicrobials using facility, prescribing, and patient indicators.
3. To identify two for each of facility, patient and prescribing indicators that increase irrational use of antimicrobials.
4. To map-out health facilities with irrational use (irrational use above WHO set percentage of irrational use) of antimicrobial.

## Methods

### Study Aim

This study seeks to identify shortcomings and areas for improvement in antimicrobial use practices. The insights gained from this analysis will be invaluable in guiding the development and implementation of targeted interventions aimed at optimizing antimicrobial usage, combating AMR, and ultimately enhancing patient care outcomes. Additionally, the study will provide a basis for formulation of key policies and intensification of regulations in antimicrobial use.

### Study Design

This will be a multiple cross-sectional study design. The study will collect primary data on prescribing indicator, facility indicator and patient indicator as per inclusion and exclusion criteria.

### Study Setting and sites

The study will collect data from CHAM health facilities. These health facilities are located in 27 of 28 political districts of Malawi (**Figure 1**). These health facilities are evenly distributed in the throughout country, with the majority located at an average distance of over eight kilometres from the nearest health facility (be it CHAM or State’s owned). Ninety-percent. Ninety percent of these health facilities are located in rural and hard-to-reach areas.

**Figure 1:**
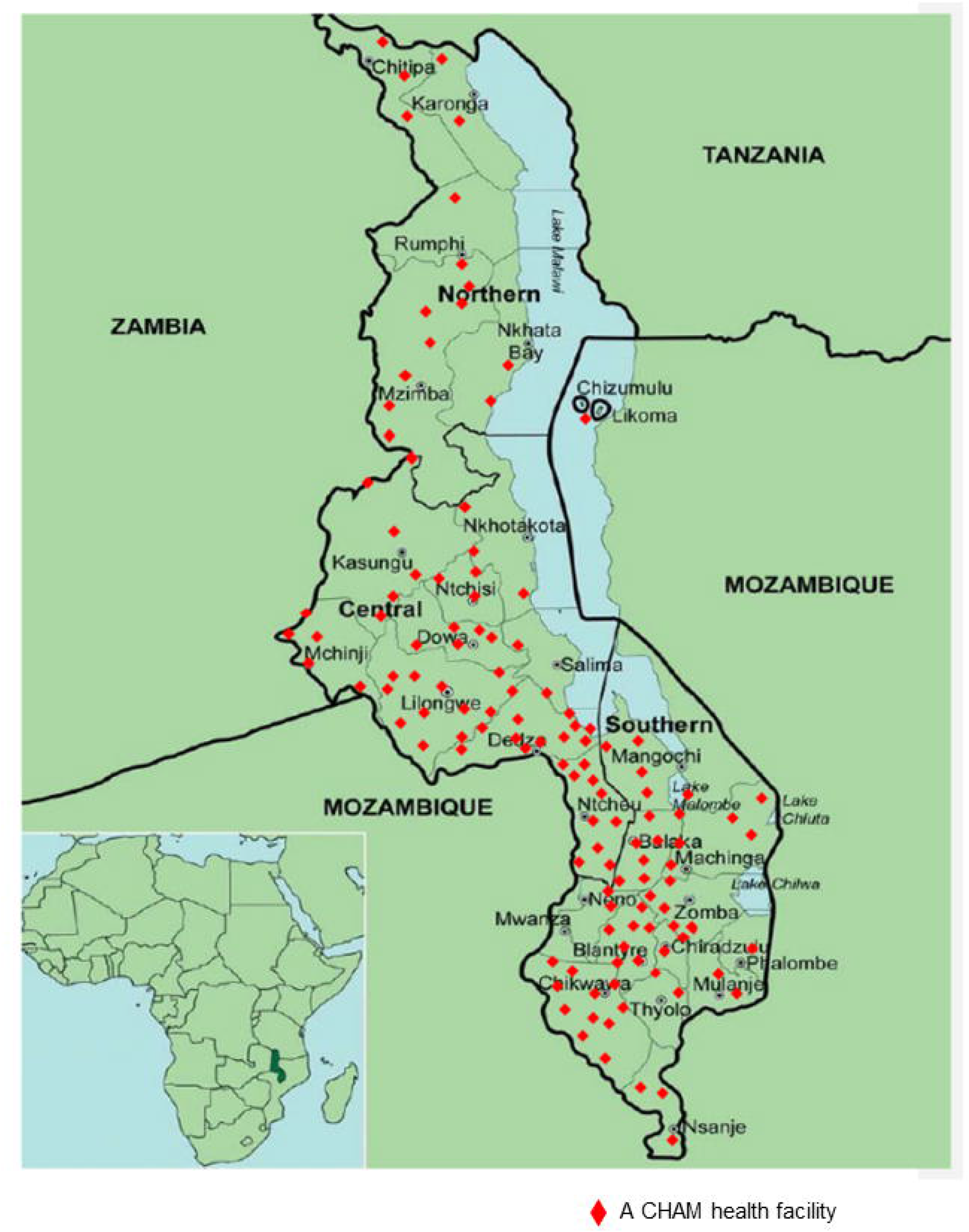
CHAM health facilities distributed across Malawi including facilities at Likoma Island.

The study will be coordinated from CHAM Secretariat in Lilongwe, the Capital City of Malawi. Additionally, data cleaning, analysis and dissemination will be planned and executed from the Secretariat.

### Participants Characteristics (Sample Size and Selection)

The study will collect data from the 194 CHAM health facilities for facility indicator. As for prescribing indicator, the sample size is 3,104 prescriptions, 16 per health facility. The 3,000 prescriptions are as per World Health Organisation (WHO) recommendation for a cross-sectional study of a prescribing indicator study (5) and the 104 are for withdrawal of consent, incomplete responses, and closure of the facilities as well as collecting data from 16 prescriptions per facility. The 3,104 prescriptions will provide enough data to determine extent of antimicrobial use and detect irrational prescribing of antimicrobial in CHAM health facilities.Patient indicator sample size is 275 patients as per cross-sectional study formula (6) with assumptions of standard normal variate (Z) of 1.96, expected proportion from previous study of 0.763 (7) and precision of 5%. The overage of 113 patients for patient indicator for withdrawal of consent, closure of facility and incomplete data, and to collect data from 2 patients per facility. This leads to a total of 388 patients for patient indicator data.

The study will collect data from all facilities for facility indicator data while systematic random sampling will be implemented for prescribing and patient indicators’ data. For prescription indicator data, each ward will contribute an equal number of participants, chosen from every even number of patient until the 16^th^ prescriptions. The patient indicator data will be collected from every odd number of the prescribing indicator data. Subsequent patient indicator data collection will utilize a new set of odd numbers for each facility. For example, if 1 and 3 were initially selected, the next round would include 5 and 7, then 9 and 11, restarting the cycle upon reaching the 15^th^ number out of the 16 prescriptions per facility.

### Data collection

Two trained personnel will collect data for each facility; pharmacy profession will collect prescribing and facility indicators data while a clinician will collect patient data. The study will use three online questionnaires, one for each indicator, on KoboToolbox v2021.2.4. Facility indicator data will be the first to collect. This will followed by simultaneous collection of patient and prescribing indicators’ data.

The study will collect data from a facility which will be opened and operational on the day of data collection (all facilities closed or suspended services during data collection). Additionally, prescribing indicator data will come from legible patient files less than a month and for patients aged 6 and above. The age for prescribing indicator will be inclusion for patient indicators’ data. Patients in intensive care will be excluded from the study.

### Study Outcome

For prescribing indicator, the study will provide the extent of antimicrobial use through percentage of hospitalization with one or more antimicrobial used and average number of antimicrobials prescribed per hospitalization in which antimicrobials were prescribed. Furthermore, the findings will include average cost of antimicrobials prescribed per hospitalization, average duration of prescribed antimicrobial treatment for intensity of exposure to antimicrobial, percentage of antimicrobials prescribed consistent with the hospital formulary list and by generic name to measure the degree of conformity to national prescribing policies, and average number of drug encounter to measure the degree of polypharmacy.

The patient indicator data outcome are percentage of prescribed antimicrobials actually administered or dispensed to a patient, average consultation and dispensing time, percentage of medicines actually dispensed, percentage of medicines adequately labelled, and percentage of patients’ correct dose. The facility indicator will provided existence of Standard Treatment Guidelines (STG) and Essential Medicines List (EML), availability of key antimicrobials in the hospital, average number of days that a set of key antimicrobials is out of stock, and percentage antimicrobial expenditure of hospital medicines cost.

Overall, the study will provide prevalence of irrational use of antimicrobial, main drivers of irrational antimicrobial use, and geographical distribution of irrational use of antimicrobials in CHAM health facilities.

### Data Management Plan

This data from KoboToolbox v2021.2.4 will be downloaded to Microsoft Excel Version 2016, cleaned and coded. The data will be stored on a password-protected computer, and access will be restricted to only study team, and relevant authority under full authorization of the PI. The data will not be connected to patients as numbers and letters will be used to identify participants. Data will be archived for any future research and participants will assent and consent to the future use of data for research purposes.

### Safety Plan

This is an observational study hence there are no direct safety concerns from the study approach. However, data collecting team will call upon clinical team to attend to any patient requiring clinical attention while responding to the study.

### Data Type and Statistical Analysis

Statistical analysis will be performed using STATA SE version 14. The data analysis will include frequencies, mean, standard deviation (SD) and percentage to describe data distribution and spread. Additionally, the study will use one-way analysis of variance (ANOVA) for numerical data and Pearson’s chi-squared or fisher’s exact (depending on the cell number) tests for categorical data, to check for association among different variables. P-value less than 0.05 will be considered statistically significant at 95% confidence interval for all data.

### Ethical Considerations and Declarations

The protocol has been approved by College of Medicine Research Ethics Committee (COMREC) on 15^th^ May 2024 [protocol number P.04/24-0651]. Prescriptions, patients and health facilities will be identified by numbers. Consent will be sought from adults while assent will be obtained for children. Participation in the study will be voluntary. Additionally, before commencing data collection, data collecting team will get approval from each health facility management team. The study team will also get approvals from the facility in-charges before commencement of the study.

### Study Status and Timeline

The study will run from January 2024 to December 2025. Currently, the implementation is at preparatory stage which will be followed by pre-testing the questionnaires then data collection up-to data dissemination (**Figure 2**).

**Figure 2:**
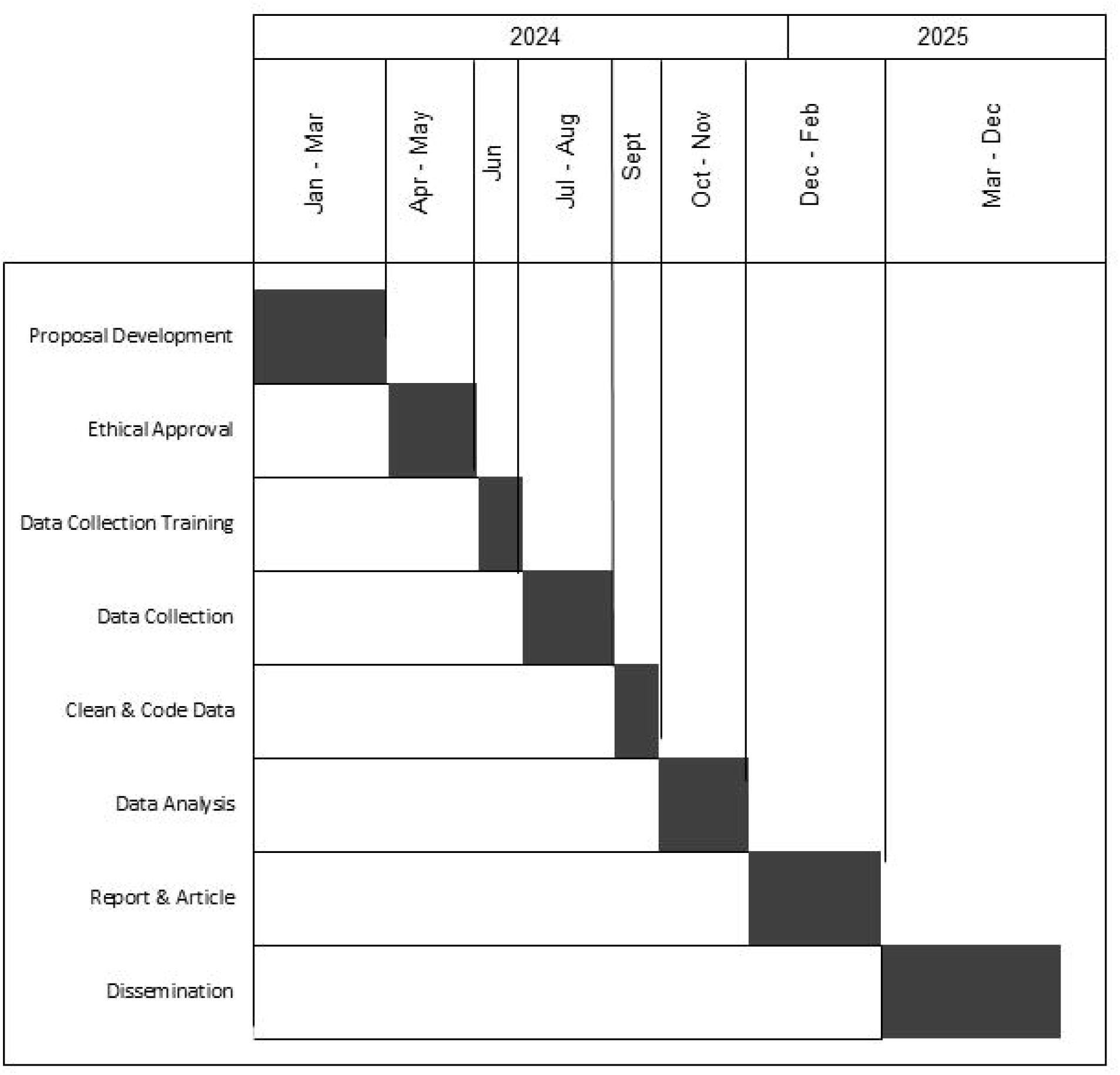
Gantt chart of the study timelines.

## Discussion

### Study Limitations and Challenges

The study will involve self-reporting and review of documents which might introduce errors because of the knowledge level of the reporter or the quality of documentation, possibly affecting the accuracy and quality of collected data. To overcome this, multiple data sources such as patients, patient file, health passport and input from the clinical team will be used. Additionally, data from patient files and health passport may be incomplete or inaccurate because of unclear handwriting or poorly recorded information. This will be addressed by excluding prescriptions with poor handwriting or incomplete records.

Logistics challenge is a concern for CHAM health facilities which are mostly in remote areas. This will be addressed by effective planning and communication, using a combination of multiple communication channels. Additionally, the study involves managing and analyzing large volume of data. This will be mitigated by thoroughly training data reporters to ensure collected data requires minimal cleaning, and by using KoboToobox to reduce most of the manual work required for data entry.

### Dissemination plan

The findings will be shared to all CHAM facilities at CHAM Annual General Meeting (AGM), the MOH AMR Sub technique working group (TWG) and Kamuzu University of Health Sciences (KUHES) Research Dissemination Conference. Additionally, the study will be presented in other local and international conferences on AMR. Finally, the study findings will be considered for publication in peer reviewed journals.

### Study Amendment and Termination

Study alterations will be done in full consultation with all study investigators with approval from ethical oversight committee. In the end, we will provide reports and communication to all key stakeholders in the project following termination of the study.

## Authors Contribution

ECC will lead and coordinate all activities as the chief investigator. DN and FC will handle all write-ups, project planning and implementation. SO, PK, SM will review, collect and analyse, and draft report and journal article. PM, EM, EK,CM, and HM will serve as overall study supervisors, providing leadership, reviews and coordinating resources for the study.

## Acknowledgements

We thank CHAM’s management for authorizing the implementation of this study in the organisation’s health facilities.

## Support Information

Ethical approvals from Malawi’s COMREC, and authorization letter from CHAM to conduct the study in the organisation’s health facilities.

## Notes

**Competing Interest** The authors declare no conflict of interest.

### Competing Interest Statement

The authors have declared no competing interest.

### Funding Statement

The author(s) received no specific funding for this work.

### Author Declarations

INSTITUTIONS OF AFFILIATION: The study is affiliated to three institutions and these are (1) Christian Health Association of Malawi (CHAM), (2) Malawi-Liverpool Wellcome Programme (MLW), and (3) Kamuzu University of Health Sciences (KUHES; formerly University of Malawi, College of Medicine). ETHICS COMMITTEE/IRB: The study received ethical approval from College of Medicine Research Ethics Committee (COMREC). The ethics approval number is P.04/24-0651.

